# MULTIMODHAL, a multicenter phase 3 randomized controlled trial: fMRI-based symptom capture for guiding rTMS treatment of drug-resistant hallucinations in patients with schizophrenia

**DOI:** 10.64898/2026.01.13.26344004

**Authors:** Renaud Jardri, Pierre Yger, Zaineb Amor, Marion Plaze, Ali Amad, David Roman, Sébastien Szaffarczyk, Stéphanie Lefebvre, Delphine Pins, Macarena Cuenca, Guillaume Coudriet, Arnaud Cachia, Julien Labreuche, Emeline Cailliau, Christine Delmaire, Olivier Outteryck, Renaud Lopes, Jean-Pierre Pruvo, Myriam Edjlali-Goujon, Catherine Oppenheim, Maxime Bubrovszky, Guillaume Vaiva, Pierre Thomas, the MULTIMODHAL Study Group, Philippe Domenech, Arnaud Leroy

## Abstract

**Background:** Auditory–verbal hallucinations (AVHs) often persist despite optimized antipsychotic regimens. While conventional low-frequency repetitive transcranial magnetic stimulation (rTMS) at the T3P3 scalp site shows limited efficacy due to interindividual variability in AVH-related brain networks, the MULTIMODHAL trial assessed whether fMRI-guided rTMS is superior to conventional targeting for drug-resistant AVHs.

**Methods:** This multicenter, double-blind, controlled trial enrolled 73 adults with drug-resistant AVHs, randomized to receive 10 sessions of active 1-Hz rTMS via either individualized fMRI-guided neuronavigation or conventional T3P3 localization. The primary outcome was the change in the Auditory Hallucination Rating Scale (AHRS) at one month post-treatment. Secondary outcomes included: patient-reported intensity and frequency, alternative severity scales, and AHRS follow-up at 3, 6, and 12 months. Mechanistic analyses assessed rTMS-induced electrical fields modeling, and baseline resting-state functional connectivity profiles.

**Results:** fMRI-guided rTMS yielded a greater reduction in AHRS scores at one month (mean difference, -5.43; 95% CI, -8.92 to -1.94), with sustained benefits at three and six months. The number-needed-to-treat for neuro-guided rTMS was 3.5. Clinical response was associated with greater E-field overlap with AVH-related networks rather than energy deposition or baseline functional connectivity. Both interventions were well-tolerated.

**Conclusions:** fMRI-guided neuronavigation significantly enhances rTMS efficacy for drug-resistant AVHs, with sustained benefits up to six months and a favorable safety profile. These findings emphasize that precise network engagement is more critical for neuromodulation success than stimulation intensity or resting-state connectivity. Study limitations include sample-size and progressive dropout after three months, warranting research on response predictors and maintenance strategies.

## Introduction

Schizophrenia (SCZ) affects 0.5 to 1% of the global population, featuring psychotic symptoms, motivational deficits, and cognitive impairments [1]. Auditory-verbal hallucinations (AVHs) are among its most disabling manifestations, commonly associated with functional disability and increased suicide risk [2]. Despite pharmacological advances, up to 40% of patients still experience persistent AVHs after multiple rounds of antipsychotic medication [3, 4, 5], highlighting the need for innovative therapeutic strategies.

Early non-invasive brain stimulation studies offered hope for drug-resistant AVHs [6]. Among these approaches is low-frequency repetitive Transcranial Magnetic Stimulation (rTMS), which applies focal magnetic pulses through a scalp-mounted coil to reduce cortical excitability. Initial clinical trials targeted the left temporo-parietal junction, an area known to be involved in both speech processing and AVH generation[7, 8]. Coil placement (and repositioning across sessions) relied on the international 10-20 system, using the “T3P3” site marked on an elastic cap fitted to the patient’s head [9]. This pragmatic yet imprecise approach has been used for years as an add-on therapy for drug-resistant AVHs [10, 11, 12, 13]. However, its estimated effect size was later revised downward [14], limiting its impact on daily clinical practice.

More recent trials have explored alternative paradigms to maximize efficiency, including bilateral stimulation [15], but a key limitation of “standard” rTMS remains the accuracy of coil placement over a biologically-relevant target, especially given the large between-subject variability in the hyperactive brain regions associated with the occurrences of AVHs. Although symptom-capture studies have confirmed a role for the temporo-parietal junction in AVH generation [16, 17, 18], they have also revealed modality-dependent activation distributed across complex association networks [19, 20, 21]. To further complicate matters, the temporo-parietal junction consists of two-to-three distinct functional subregions involved in different cognitive processes (ranging from attention to self-other distinction), which may not overlap functionally [22, 23].

Hence, neuronavigating rTMS to the most accessible cortical hubs within AVH-related networks may be critical for maximizing treatment efficacy [24, 25, 26]. Progress was previously hindered by the inherent limitations of motor-based symptom-capture fMRI [27] and the longstanding lack of a reliable procedure for individualizing rTMS targets [28]. These technical barriers were recently overcome by validating a reproducible and reliable method for identifying AVH-related brain clusters at the individual level [19, 29, 30], even in highly symptomatic SCZ patients. Together, these developments enable personalized cortical targets based on pre-TMS fMRI AVH-capture scans.

Because the mechanistic relationship between such functional biomarkers and AVHs was supported only by neuronavigated rTMS case reports [31, 32, 33], we conducted the first randomized phase-3 controlled trial directly comparing fMRI-guided rTMS with conventional T3P3 cap-targeting in 73 schizophrenia patients with drug-resistant AVHs. The primary endpoint evaluated changes in AVH severity using the Auditory Hallucination Rating Scale (AHRS) one month post-treatment, with follow-ups extending to 12 months.

## Methods

### Experimental Design

MULTIMODHAL was a phase 3, multicenter, prospective, randomized, double-blind controlled trial designed to test the superiority of neuronavigated rTMS over T3P3 rTMS in reducing AVH severity in SCZ patients. Participant screening, treatment, safety and follow-up were conducted from June 2011 to February, 2022 at two centers in France (CHU Lille and GHU Paris, Psychiatrie & Neurosciences). After undergoing an fMRI scan and confirming that a personalized brain target could be successfully identified, patients were randomized into two arms to receive active non-invasive brain stimulation. Regular clinical assessments were scheduled during the 12-month follow-up period. The trial complied with the International Council for Harmonization Good Clinical Practice guidelines, received national ethics approval (CPP Nord-Ouest, registration # 2009-A00842-55), and was registered at ClinicalTrial.gov. The final plan of analysis, developed in accordance with the SPIRIT guidelines, is provided in the Supplementary Protocol section. Independent data and safety monitoring were ensured by the research sponsor. The funding agency played no role in the design execution of the trial, analysis or data reporting.

### Targeted Population

Eligible patients were aged 13 to 60 years old, with a confirmed diagnosis of schizophrenia according to the DSM-IV-TR criteria, and were experiencing drug-resistant AVHs (i.e., persistent hallucinations after two consecutive lines of antipsychotic medications at an adequate dosage for at least 8 weeks). Although the protocol permitted the inclusion of minors, no participants under 18 were ultimately randomized. To control for brain asymmetry in the targeted brain areas [34], only right-handed participants were included. Additional inclusion criteria involved the absence of comorbid neurological disorders or current substance abuse and the use of stable antipsychotic medication for at least 30 days prior to rTMS (with doses maintained without modification through the 3 first months evaluation window). Participants who were pregnant, had contraindication to MRI scanning or magnetic stimulation, had claustrophobia and lacked social insurance were excluded. All of the participants provided oral and written informed consent prior to enrollment.

### Randomization & Blinding

Participants were randomly assigned in a 1:1 ratio to receive either neuronavigated or standard T3P3 rTMS via sealed envelopes, using a computer-generated sequence (block sizes of six, stratified by center). To preserve the per-protocol sample size, participants who were randomized but discontinued intervention before receiving rTMS were replaced. Participants, clinicians and investigators were blinded to group allocation and remained unaware of the grouping throughout the trial, except for the TMS operator, who did not participate in any clinical assessments. For the participants, blinding was maintained at each session using an elastic cap (used to define the T3P3 site in the standard group), paired with an active neuronavigator mounted on the TMS coil (used in the neuro-guided group). The success of blinding was assessed post-treatment via *Bang’s Blinding Index* [35].

### Trial Interventions

Both groups received active rTMS, as prior trials and meta-analyses have demonstrated the superiority of active rTMS over sham stimulation in reducing AVH [10, 36, 11]. Treatment comprised ten sessions of 1200 pulses each, delivered twice daily at 1-Hz over 5 consecutive days (referred to as the “intervention period”) [37]. The two daily sessions were separated by a strict 5-hour interval. Stimulation intensity was set at 100% of the resting motor threshold (RMT, visually determined during the first session by tracking contractions of the abductor pollicis brevis muscle), but could be reduced stepwise by 5% increments to a minimum of 70% for comfort. Once established, the stimulation amplitude was maintained for all 10 sessions. Treatments were delivered using a MagPro X100 stimulator equipped with a B65 fluid-cooled coil (MagVenture). Depending on randomization, coil placement was either (i) neuronavigated to a personalized brain target based on pre-TMS AVH-capture fMRI scans [19, 30] (the “neuro-guided group”, see details in the Supplementary Methods), or (ii) located at the T3P3 scalp location using the 10-20 cap targeting method [9] (the “standard group”). Safety assessments included systematic monitoring for adverse events during or after the rTMS sessions.

### Outcome Measures

#### Choice of the primary measure

The protocol originally anticipated a broader cohort of multisensory hallucinations [38, 39], for which no validated instrument was available at that time. Consequently, we initially registered the use of self-reported measures via *Visual Analogue Scales* (VAS, a general measure insensitive to specific hallucination modalities) as the primary outcome. However, during the initial screening and enrollment phases, it became apparent that approximately 90% of eligible participants presenting with drug-resistant phenomena primarily experienced AVHs. To prevent the trial from being insensitive to nuanced changes in AVH topography (such as frequency, loudness or related distress), the analytical focus was shifted to an AVH-specific metric at 1-month (see also **Sample-Size Calculation**). The *Auditory Hallucination Rating Scale* (AHRS) was designated as the primary outcome prior to unblinding, with the original VAS scores retained as key secondary metrics (see **Alternative Measures of Severity**). To demonstrate the negligible impact of this mid-trial adaptation, results for both the seminal and final primary endpoints are presented Fig.1A.

**Figure 1.**
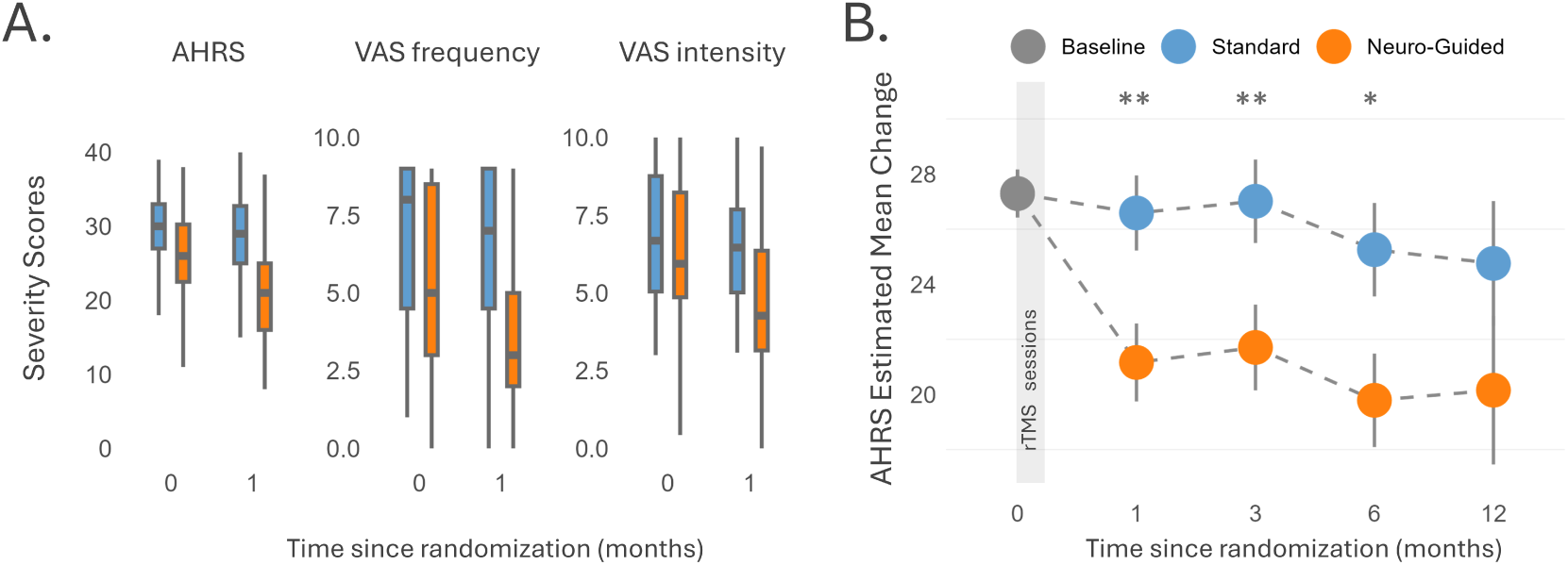
Efficacy analysis and longitudinal outcome of rTMS for Auditory-Verbal Hallucinations (AVH). Colors represent groups: standard (blue) and neuro-guided (orange). **(A)** Box-plots of raw-data at baseline (0) and 1-month follow-up (1), for Auditory Hallucination Rating Scale (AHRS, primary end-point), and patient-reported VAS for frequency and intensity (key secondary outcomes). **(B)** Constrained Longitudinal Data Analysis illustrating between-group adjusted estimated mean differences on Auditory Hallucination Rating Scale (AHRS) scores at 1, 3, 6 and 12 months post-rTMS treatment. Error bars represent standard errors. * p<0.05, ** p<0.01.

### Primary and secondary endpoints

Participants were evaluated at screening and baseline, as well as at 1, 3, 6, and 12 months after treatment. The **primary endpoint** was the change in AVH severity from baseline, assessed by the AHRS score at 1 month of rTMS treatment initiation (i.e., the difference between the AHRS score at 1 month and that at baseline).

The following secondary endpoints were analyzed:

- **Key secondary outcome**: a change from baseline in hallucination severity based on *Patient Reported Outcome* (P.R.O.) measures utilizing Visual Analogue Scales (VAS) for frequency and intensity of the main hallucinatory sensory modality, retaining the capacity to assess rTMS effects on potential non-auditory hallucinations;
- **Alternative measures of severity**: a change from baseline in AVH severity based on alternative measures, such as *Positive And Negative Syndrome Scale* (PANSS) score and sub-scores [40, 41], *Clinical Global Impression* (CGI) improvement score, and *Global Assessment of Functioning* (GAF) score, at 1 month of rTMS session initiation (i.e., the difference in score values assessed at 1 month and baseline, except for the CGI improvement score which was only assessed at 1 month);
- **Longitudinal measures of severity**: a change from baseline in hallucination severity assessed via AHRS scores at 3, 6, and 12 months after rTMS treatment;
- **Modeling of rTMS-induced cortical electric fields**: (i) comparison between responders and non-responders regarding individualized AVH-network engagement by the simulated E-field, including cumulative unsigned IC weights within the stimulation area, total E-field extent and total deposited energy; and (ii) comparison between responders and non-responders of baseline resting-state functional connectivity profiles within the simulated E-field.

### Statistical Methods

#### Sample Size Calculation

Based on meta-analyses of the effects of T3P3 rTMS available at the time [10, 36, 11], we planned to randomize 70 participants (35 per group), to achieve 80% power in detecting a between-group difference of 1.5 points on the VAS scores at 1-month (standard deviation: 2 points, effect-size: 0.75), using a two-sided t-test with a 0.05 significance level and a 20% dropout rate. Crucially, substituting the AHRS as the primary endpoint (See Choice of the primary measure), with an estimated standard deviation of 5 points, still guaranteed sufficient power to detect a mean difference of 3.75 points between groups. Analyses of the P.R.O.s are now presented in the secondary efficacy outcomes section.

### Primary & Secondary Efficacy Outcomes

Primary efficacy analyses were conducted on the full analysis set, which included all of the randomized patients regardless of eligibility violations or protocol deviations, who were analyzed according to their assigned treatment-group (referred to as **intention-to-treat** or **ITT**). The change from baseline in hallucination severity (as assessed using the primary outcome (AHRS score) as well as secondary P.R.Os), was compared between groups by using a **constrained longitudinal data analysis** (cLDA) [42, 43]. This approach estimated baseline-adjusted mean differences in follow-up changes in AVH severity for neuro-guided versus standard rTMS, with 95% confidence intervals (CIs) reported as the treatment effect-size. In cLDA, both baseline and post-baseline values were modeled as dependent variables using a linear mixed model for longitudinal data, with an unstructured covariance pattern model and the baseline means constrained to be equal across the two treatment groups. This approach allows to account for baseline differences within the longitudinal model and uses all available baseline and post-baseline observations, with treatment effects estimated from the time-by-treatment-group interaction. For outcome measures where cLDA residuals deviated from normality (i.e., PANSS P3-item and VAS frequency), absolute changes from baseline to the follow-up time-points were calculated. These data were compared between groups by using a non-parametric analysis of covariance (ANCOVA), adjusted for baseline values. Treatment effect-sizes were expressed as standardized mean differences, with 95%CIs calculated for the rank-transformed data.

### Sensitivity Analyses

Two **sensitivity analyses** were pre-specified: (i) in the first analysis, missing values were handled via multiple imputations (20 imputations) under the missing-at-random assumption [44] by using treatment groups and key baseline characteristics (Table 1). Imputation employed fully conditional specification with predictive mean matching for quantitative variables and logistic regression (binomial, ordinal or multinomial for categorical variables); (ii) in the second one, analysis was restricted to the per-protocol population (patients who received their allocated intervention). Treatment-effect estimates from the imputed datasets were combined using Rubin’s rules [45].

**Table 1.** Baseline Demographic and Clinical Characteristics of Participants in the MULTIMODHAL Randomized Trial.

|  | Neuro-Guided<br>(n=36) | Standard<br>(n=37) |
| --- | --- | --- |
| Age (in years) | 37 ±10 | 37 ±11 |
| Sex, female | 13/36 (36.1) | 16/37 (43.2) |
| Educational Level: |  |  |
| Low education (ISCED 1,2): | 24/35 (68.6) | 29/37 (78.4) |
| Medium education (ISCED 3,4): | 11/35 (31.4) | 8/37 (21.6) |
| Age at Onset (years) | 22 ±8 [n=27] | 21 ±7 [n=31] |
| AHRS score | 25 ±8 [n=35] | 29 ±6 |
| PANSS total score | 81 ±23 | 76 ±14 |
| PANSS positive factor | 18 ±5 | 16 ±4 |
| PANSS negative factor | 21 ±8 | 21 ±6 |
| PANSS disorganized factor | 20 ±7 | 18 ±5 |
| PANSS excitation factor | 5 (4 to 8) | 5 (4 to 6) |
| PANSS depression factor | 13 ±5 | 13 ±4 |
| PANSS P3-item | 5 (5 to 6) | 6 (5 to 6) |
| VAS frequency | 5 (3 to 9) [n=35] | 8 (4 to 9) [n=35] |
| VAS intensity | 6.2 ±2.6 [n=35] | 6.9 ±2.1 [n=35] |
| GAF score | 43 ±15 | 47 ±13 |
| Olanzapine Equivalent Dose (mg) | 32 (24 to 56) | 40 (23 to 52) |
| Diazepam Equivalent Dose (mg) | 8 (0 to 38) | 10 (0 to 27) |
| Treatment Intensity (% of Motor Threshold) | 88.8 (70 to 100) | 90.5 (80 to 100) [n=35] |

Values are no./total no. (%), means ± standard deviation or median (25th to 75th percentile); in cases of missing data, the number of available cases has been reported in bracket [n]. Educational levels are provided in reference to the *International Standard Classification of Education* (ISCED). AHRS stands for *Auditory Hallucination Rating Scale*. PANSS stands for *Positive And Negative Syndrome Scale*. GAF stands for *Global Assessment of Functioning* scale. CGI stands for *Clinical Global Impression* scale.

Two **post-hoc sensitivity analyses** were also conducted. The first unplanned sensitivity analysis evaluated the robustness of the longitudinal findings (AHRS score changes) under two distinct missing-data scenarios. The imputation approach was determined according to the observed reason for missingness: (i) the last available value was carried forward for patients considered true dropouts, whereas (ii) the baseline value was used for patients who received a therapeutic release. The second explored the impact of the extended recruitment period (2011 - 2022) by performing a leave-one-out sensitivity analysis, dividing the total timeframe into five smaller sub-periods and assessing the primary outcome while excluding each sub-period one by one.

All of the statistical tests were two-sided, with a significance threshold of p<0.05. Due to the risk of type I error from multiple comparisons, secondary outcome analyses should be interpreted as exploratory. These analyses were performed at the Statistics, Economic Evaluation and Data Management Unit - SEED, Lille University Hospital, France, using the SAS software (Version 9.4. SAS Institute Inc, Cary, NC, USA).

### Quantitative assessment of the electrical field - AVH network interaction

First, we retrospectively simulated the electrical field (E-field) induced by rTMS [46] for each patient, based on coil location, orientation and the stimulation amplitude (percentage of the resting motor threshold, RMT%) recorded during therapeutic sessions (see details in the **Supplementary Methods**). For all of the subsequent analyses, the **rTMS stimulation area** for each individual was defined as the 15% RMT-thresholded E-field. To assess the impact of rTMS targeting accuracy, we computed three additional metrics: (a) *the extent of the E-field*, which was quantified as the number of MRI voxels in the patient’s cortex, (b) *the extent of AVH network engagement*, which was quantified as the sum of the unsigned AVH-voxel weights falling into the stimulation area, and (c) *the total energy deposited* within the stimulation site. These metrics were compared between responders and non-responders using non-parametric permutation-based logistic regression with 10,000 label permutations.

Second, we explored the E-field functional connectivity profile in responders versus non-responders. After normalization, an atlas-based parcellation was applied to individual MR images [47], and baseline resting-state Functional Connectivity (FC) was computed by using each individual rTMS-induced E-field as the primary seed. A spectral K-means algorithm, with the optimal number of clusters determined by silhouette index scores [48]. Tgrouped elements with similar features into the same cluster. Detailed pre-processing and analysis steps are reported in the **Supplementary Methods**. The results were projected onto MNI-standardized cortical surfaces (Glasser Atlas). Group-level FC profiles were first displayed as polar plots for the 20 top-parcels common to both groups, allowing a qualitative assessment of connectivity differences. Second, we quantitatively assessed how (i) the number of voxels within the identified clusters per subject, and (ii) the patient-specific overlap between these clusters and the AVH network, accounted for the response classification, using a non-parametric permutation-based logistic regression with 10,000 label permutations.

## Results

### Trial Participants

Of 157 patients screened in person, 73 met inclusion criteria and were randomized (none were under 18 y.o.). Specifically, 36 were allocated to the “neuro-guided rTMS” group, and 37 to the “standard rTMS” group. The two groups exhibited similar baseline characteristics (see Table 1). All but three participants completed the full treatment and immediate post-treatment assessment (one discontinued after one rTMS session; two had non-contributive symptom-capture fMRI scans). Both arms received comparable stimulation doses (88.8% versus 90.5% of the RMT). Per protocol, participants whose AVHs returned to baseline severity after 3 months could withdraw to pursue alternative treatments (n=44, 65% in the control group and 60% in the neuro-guided rTMS group). This attrition warrants caution when interpreting results beyond this time-point. No withdrawals were due to adverse events (see Safety section and the CONSORT flow-diagram Supplementary Figure 4). The blinding procedure appeared effective, as participant’s guesses about treatment group allocation yielded a Bang’s Blinding Index of 0.16 ([0–1] range, successful below 0.2).

### Efficacy Outcomes

In the **primary efficacy (ITT) analysis**, neuro-guided stimulation led to a significantly greater reduction in AHRS scores at 1-month. Patients receiving neuro-guided rTMS exhibited a mean change from baseline of -6.13 pts (95%CI, -8.68 to -3.58) versus -0.71 pts (95%CI, -3.14 to 1.73) in those receiving T3P3 cap-targeting rTMS. The baseline-adjusted mean difference was -5.43 (95%CI, -8.92 to -1.94, Table 2, Fig.1). Similar findings were obtained after missing values were addressed with multiple imputations (baseline-adjusted mean difference of -5.52, 95%IC, -8.99 to -2.04) or when the analysis was restricted to the per-protocol population (baseline adjusted mean difference of -5.72, 95%IC, -9.54 to -1.90 (see the Supplementary Tables 4,5). Leave-one-out analyses on sub-periods of inclusion yielded similar trends (see Supplementary Figure 5).

**Table 2.** Primary and Secondary Outcomes of the MULTIMODHAL trial: Primary efficacy (ITT) analysis.

|  | N | Neuro-Guided<br>(n=36) | N | Standard<br>(n=37) | Effect Size (95% CI) | p-Value |
| --- | --- | --- | --- | --- | --- | --- |
| **Primary Outcome (AHRs Score)** |  |  |  |  |  |  |
| 0 (baseline) | 35 | 25 ±8 | 37 | 29 ±6 |  |  |
| 1 Month | 31 | 20 ±9 | 34 | 28 ±7 |  |  |
| 3 Months | 23 | 21 ±9 | 24 | 27 ±8 |  |  |
| 6 Months | 21 | 18 ±10 | 20 | 25 ±8 |  |  |
| 12 Months | 10 | 17.8 ±10.9 | 14 | 24 ±6 |  |  |
| diff. 1-0 |  | -6.1 (-8.6 to -3.6) |  | -0.7 (-3.1 to 1.8) | -5.4 (-8.9 to -2.0) | **0.003** |
| diff. 3-0 |  | -5.6 (-8.3 to -2.9) |  | -0.3 (-2.9 to 2.4) | -5.3 (-9.1 to -1.6) | **0.007** |
| diff. 6-0 |  | -7.5 (-10.5 to -4.5) |  | -2.0 (-5.0 to 1.0) | -5.5 (-9.7 to -1.2) | **0.014** |
| diff. 12-0 |  | -7.1 (-12.5 to -1.8) |  | -2.5 (-12.5 to -1.8) | -4.6 (-11.5 to 2.4) | 0.18 |
| **Secondary Outcomes** |  |  |  |  |  |  |
| PANSS total score |  |  |  |  |  |  |
| 0 (baseline) | 36 | 81 ±23 | 37 | 76 ±14 |  |  |
| 1 Month | 34 | 66 ±26 | 34 | 68 ±22 |  |  |
| diff. 1-0 |  | -13.6 (-20.6 to -6.7) |  | -8.6 (-15.6 to -1.7) | -5.0 (-14.7 to 4.7) | 0.31 |
| PANSS positive sub-score |  |  |  |  |  |  |
| 0 (baseline) | 36 | 18 ±5 | 37 | 16 ±4 |  |  |
| 1 Month | 33 | 15 ±5 | 34 | 16 ±4 |  |  |
| diff. 1-0 |  | -2.4 (-3.3 to -1.4) |  | -0.8 (-1.8 to 0.2) | -1.6 (-2.9 to -0.2) | **0.026** |
| PANSS P3-item |  |  |  |  |  |  |
| 0 (baseline) | 36 | 5 (5 to 6)<sup>a</sup> | 37 | 6 (5 to 6)<sup>a</sup> |  |  |
| 1 Month | 33 | 5 (4 to 5)<sup>a</sup> | 34 | 5 (5 to 6)<sup>a</sup> |  |  |
| diff. 1-0 |  | 0 (-2 to 0)<sup>a</sup> |  | 0 (-1 to 0)<sup>a</sup> | -0.4 (-0.8 to 0.1)<sup>b</sup> | 0.098 |
| VAS frequency |  |  |  |  |  |  |

Table 2 – continued from previous page
|  | N | Neuro-Guided<br>(n=36) | N | Standard<br>(n=37) | Effect Size (95% CI) | p-Value |
| --- | --- | --- | --- | --- | --- | --- |
| 0 (baseline) | 35 | 5 (3 to 9)<sup>a</sup> | 35 | 8 (4 to 9)<sup>a</sup> |  |  |
| 1 Month | 35 | 3 (2 to 5)<sup>a</sup> | 35 | 7 (4 to 9)<sup>a</sup> |  |  |
| diff. 1–0 |  | –1 (–3 to 0)<sup>a</sup> |  | 0 (–1 to 0)<sup>a</sup> | –0.5 (–1.0 to –0.1)<sup>b</sup> | **0.020** |
| VAS intensity |  |  |  |  |  |  |
| 0 (baseline) | 35 | 6.2 ±2.6 | 35 | 6.9 ±2.1 |  |  |
| 1 Month | 35 | 4.5 ±2.5 | 35 | 6.4 ±2.3 |  |  |
| diff. 1–0 |  | –1.9 (–2.6 to –1.2) |  | –0.3 (–1.0 to 0.4) | –1.6 (–2.5 to –0.7) | **0.002** |
| GAF score |  |  |  |  |  |  |
| 0 (baseline) | 36 | 43 ±15 | 37 | 47 ±13 |  |  |
| 1 Month | 33 | 47 ±14 | 34 | 46 ±14 |  |  |
| diff. 1–0 |  | 3.9 (0.5 to 7.3) |  | 0.7 (–2.5 to 4.0) | 3.2 (–1.3 to 7.8) | 0.17 |
| CGI score at 1 Month | 34 | 3 (3 to 4)<sup>a</sup> | 34 | 4 (3 to 4)<sup>a</sup> | –0.3 (–0.8 to 0.2)<sup>b</sup> | 0.15 |

**Notes:** <sup>a</sup> Median (25th to 75th percentile). <sup>b</sup> Standard mean difference on rank-transformed data.
Values are mean ± standard deviation or least-squares means (95% CI) estimated from cLDA models unless otherwise stated. Effect sizes represent mean differences (95% CI) in baseline-adjusted changes from baseline, except where noted. AHRs stands for *Auditory Hallucination Rating Scale*. PANSS stands for *Positive And Negative Syndrome Scale*. VAS stands for *Visual Analogue Scale*. GAF stands for *Global Assessment of Functioning* scale. CGI stands for *Clinical Global Impression* scale. The effect appears specific to AVH metrics. In contrast, no between-group differences were observed for broader measures of global psychopathology or functioning, suggesting that the effect of neuro-guided rTMS was relatively symptom-specific and aligned with the target definition.

As shown in Table2, the neuro-guided rTMS group also exhibited a significantly greater one-month reduction in **secondary outcomes** specific to hallucination severity, including *patient-reported outcome* (P.R.O.) metrics for AVH frequency and intensity (see also Fig.1A), and the PANSS positive sub-score as well as a trend for the P3-item. No between-group differences were observed in secondary outcomes assessed by global psychometric tools that did not specifically focus on hallucinatory experiences (e.g., 1-month changes in the PANSS total score, GAF score or CGI-I score at one month). These patterns were consistently observed across the different planned sensitivity analyses (Supplementary Tables 4,5).

### Longitudinal Analysis

Longitudinal analysis of the therapeutic effects on the primary outcome revealed sustained benefits of neuro-guided rTMS at 3 and 6 months, with mean differences of -5.31 (95%CI, -9.10 to -1.51) and -5.47 (95%CI, -9.79 to -1.15) respectively. However, this effect was no longer significant at 12 months post-treatment, with a mean difference of -4.60 (95%CI, -11.53 to 2.32, see Table2 and Fig.1B). Unplanned sensitivity analysis as well as per-protocol analyses yielded comparable effect-size estimates, while multiple imputation analyses showed lower effect-sizes at 3, 6 and 12-months (see Supplementary Tables 4,5,6).

### Additional Analyses based on Optimized Targets and Treatment Response

To further elucidate the neurobiological mechanisms underlying the efficacy of rTMS in the treatment of AVHs, we compared responders with non-responders. We followed prior definitions of clinical response to rTMS in individuals with drug-resistant AVHs and defined responders as those with a >25% decrease in basal AHRS scores [6, 49]. In the neuro-guided rTMS group, 45.2% of the participants (n=14) were classified as treatment-responders, versus 17.6% of those in the standard rTMS group (n=6) (see **Methods**). The number-needed-to-treat was determined to be 3.5 (95%CI, 2.0 to 12.4).

Post hoc E-field modeling at the rTMS stimulation site revealed greater target spatial variability in the neuro-guided group (e.g., bilateral pattern) compared to the standard group (left-lateralized, see Fig. 2C), which was consistent with the individualized targeting strategy. Responders, regard-less of the treatment arm, had a broader cortical E-field extent (t(62)=2.057, p=0.008, *β*=0.781), and greater overlap with AVH network voxels, yielding higher cumulative absolute IC weights (t(62)=2.22, p=0.02, *β*=0.820). However, the deposited energy did not differ between responders and non-responders (see Fig.3A). These results suggest that E-field overlap with the AVH network (but not intensity) may drive clinical efficacy (see also Fig.2D). This aligns with the observed inverse association between distance from T3P3 to the optimal target and AHRS reduction at 1 month in the standard group (R²=0.17, p=0.024, see Fig.2E).

**Figure 2.**
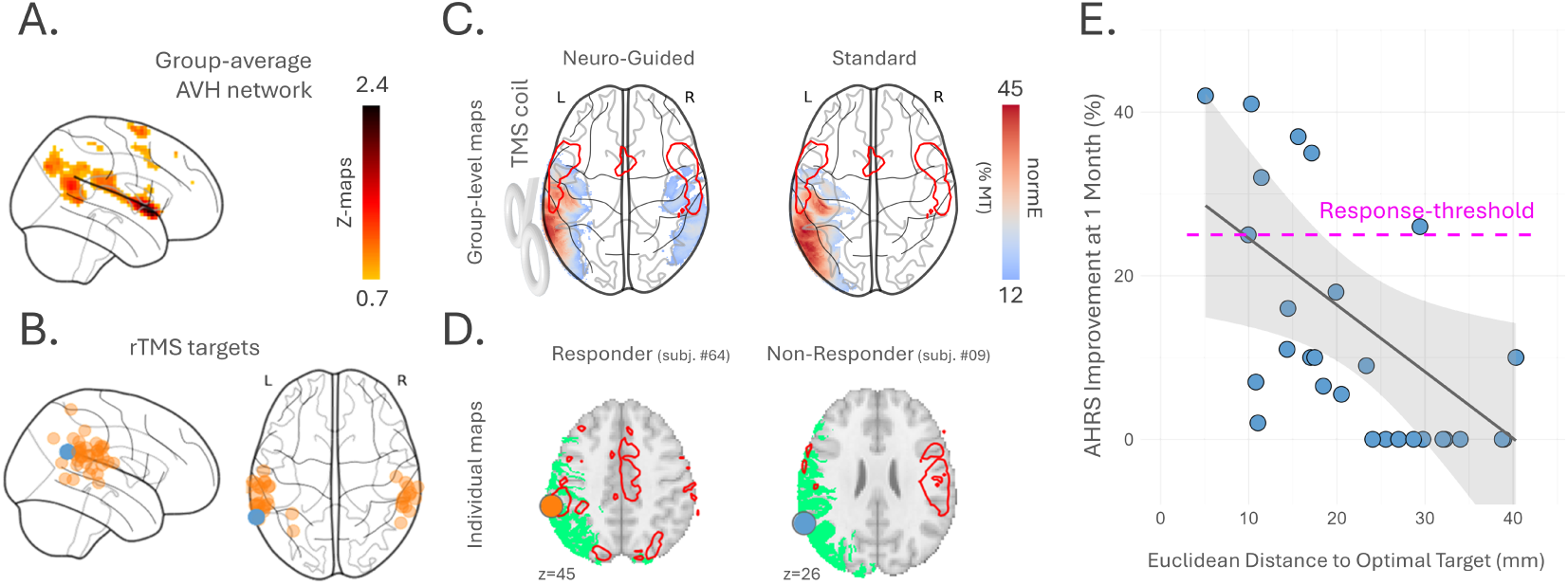
Optimal brain target definition, TMS-induced electrical fields and reduction in hallucination severity. **(A)** Lateral view of the group-average AVH networks derived from fMRI capture in the full sample. Individual maps were used to identify optimal brain targets for each participant in the neuro-guided group. **(B)** Glass-brain projections of the brain targets used in the trial, color-coded by group: standard (blue) and neuro-guided (orange). **(C)** Post-hoc computation of TMS-induced mean electrical field (E-field) distributions for participants in the neuro-guided group (left) and the standard group (right). E-fields are overlaid on the AVH network (red contours) and projected onto a glass brain. **(D)** At the individual level, the overlap between the brain target (colored disc), the cortical E-field (binarized in green) and the AVH network (red contour) is more pronounced in responders (left) than in non-responders (right, see also Figure 3). The individualized brain-based target is shown in orange (neuro-guided group), and the scalp-based T3P3 target is shown in blue (standard group). Panel D illustrates representative individual data for participant 09 and 64, respectively. **(E)** Association between the T3P3-to-optimal target distance and the reduction in hallucination severity (Auditory Hallucination Rating Scale [AHRS] scores) at 1-month in the Standard group. A greater Euclidean distance between the actual stimulation site and the fMRI-derived optimal target predicted a lesser reduction in symptoms (R²= 0.17, p=0.024). Each blue dot represents an individual participant from the standard group. Treatment-responders were defined as those achieving at least 25% clinical improvement, as indicated by the pink dashed line.

**Figure 3.**
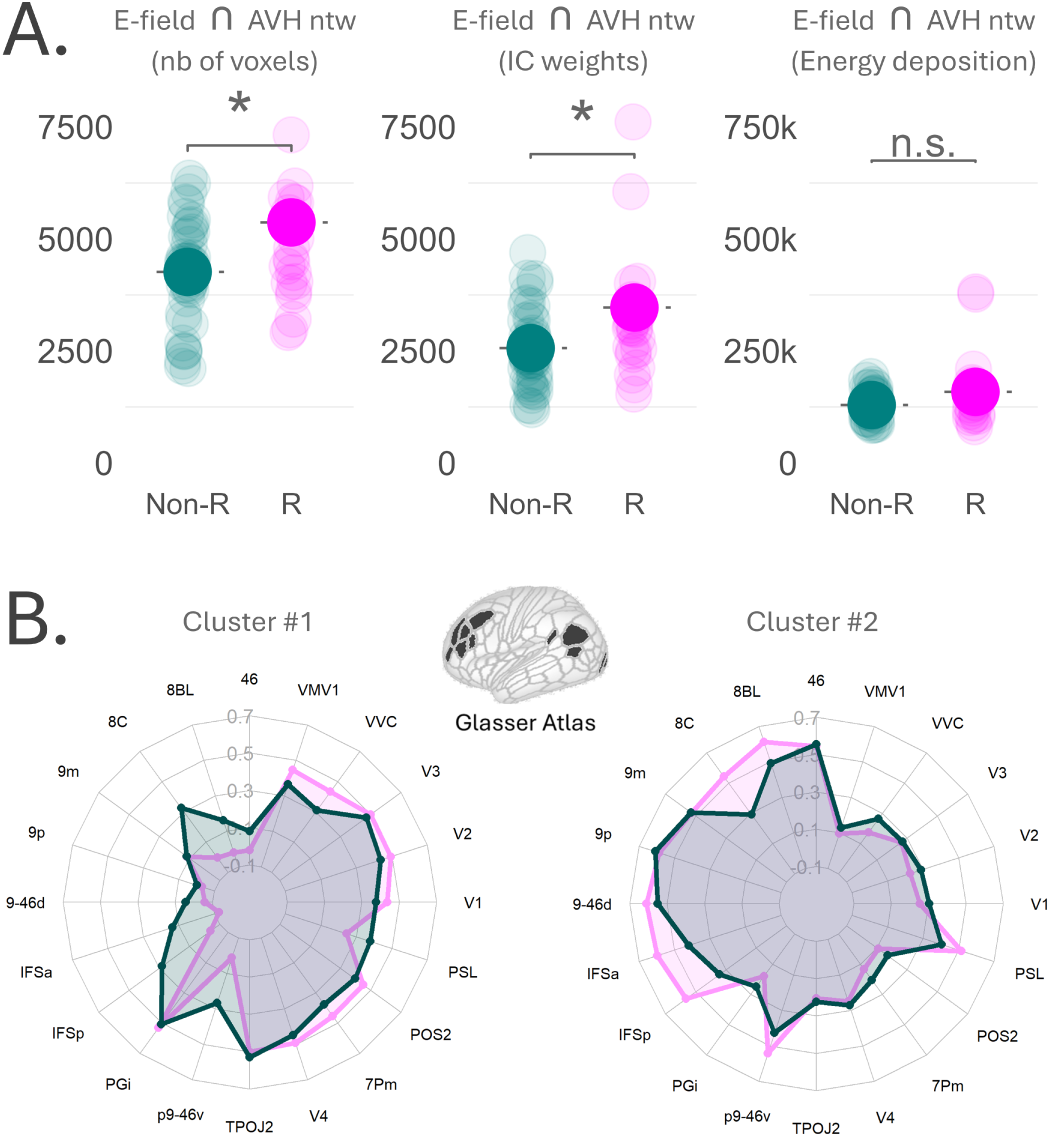
Clinical response to rTMS is driven by electrical field (E-field) interaction with the Auditory-Verbal Hallucination (AVH) network, not baseline resting-state connectivity at the stimulation site. **(A)** In responders (pink, R), the cortical E-field map shows greater spatial overlap with the AVH network than in non-responders (green, non-R). This difference is reflected in both voxel count (left panel) and functional contribution (middle panel). However, the total deposited energy within the AVH network, once recruited, is not associated with improved clinical response (right panel).**(B)** Baseline resting-state functional connectivity at the stimulation site (defined as the E-field, thresholded at 15% of the resting motor-threshold) fails to distinguish responders from non-responders. The mean connectivity strength of the top 20 parcels in cluster #1 and #2, shows no significant between-group difference (the radar plot labels correspond to the parcel abbreviations from the Glasser atlas).

Data-driven connectivity-based parcellation identified two functional clusters at the stimulation site (See Fig.3B). No significant baseline differences in response were observed between these clusters in voxel count (Cluster 1: p=0.386, Cluster 2: p=0.249) or overlap (Cluster 1: p=0.313, Cluster 2: p=0.140), suggesting resting-state intrinsic FC alone does not explain clinical response.

### Safety

No serious adverse-events occurred. Common rTMS side-effects were reported on a daily basis during the intervention period, and were similar between groups (see Table 3). The incidences of side-effect were: 11.4 - 19.4% for discomfort at stimulation site (i.e., if it occurred only during stimulation), 19.4 - 20% for post-rTMS headache (i.e., if it persisted after stimulation), and 2.8 - 2.9% for anxiety. Reported headaches resolved within hours following rTMS administration, either spontaneously or with a single administration of nonsteroidal anti-inflammatory drug.

**Table 3.** Summary of Adverse Events that occurred during the 12 Months MULTIMODHAL trial period following rTMS.

|  | Neuro-Guided<br>(n=36) | Standard<br>(n=37) |
| --- | --- | --- |
| **Overall** |  |  |
| Any A.E.: | 12/36 (33.3) | 9/35 (25.7) |
| Serious A.E.: | 0/36 (0.0) | 0/35 (0.0) |
| A.E. leading to trial discontinuation: | 0/36 (0.0) | 0/35 (0.0) |
| **Details** |  |  |
| Post-rTMS headache: | 7/36 (19.4) | 7/35 (20) |
| Discomfort at stimulation site: | 7/36 (19.4) | 4/35 (11.4) |
| Anxiety: | 1/36 (2.8) | 1/35 (2.9) |

Values are no./total no. (%). A.E. stands for Adverse Events.

## Discussion

In this 12-month, multicenter, blinded, randomized phase-3 trial, we report the superiority of neuro-guided over standard rTMS in reducing AVH severity in drug-resistant schizophrenia patients. All randomized participants (except three) completed the 5-day treatment course, with significant improvements observed at 1 month post-treatment (primary end-point) across multiple measures: AHRS, PANSS-positive subscale and patient-reported VAS scores for severity or frequency. These effects were AVH-specific, persisted through 3 and 6 months, and showed no impact on broader clinical metrics. Both standard and neuro-guided rTMS approaches were equally safe, with no difference in self-reported adverse events.

Of note, the standard (T3P3) group showed minimal symptom reduction. While active standard rTMS protocols typically yield a small therapeutic effect-size, the negligeable impact observed here underscores the severe treatment resistance of this cohort. It also suggests that delivering 1-Hz rTMS without clear personalized targeting may fail to reach the threshold dosage required to engage the AVH network effectively (i.e., in terms of the number of voxels and IC weights). While some authors addressed this issue by expanding the number of TMS targets, for instance through bilateral stimulation protocols [50, 15], our approach focuses on a precise and personalized target definition (41.6% of our neuro-guided group also received right hemisphere stimulation).

Neuronavigation is known to reduce inter-operator variability in rTMS coil placements [25]. In major depressive disorder, targeting the dorsolateral prefrontal area most functionally anticorrelated with the subgenual anterior cingulate has yielded breakthroughs [51, 52, 53]. For drug-resistant hallucinations, only three sham-controlled trials have evaluated neuronavigated rTMS, two of which showed no advantage over sham [54, 55]. These studies used different target determination methods. One relied on maximal task-evoked activation during a language task [55], which, while often overlapping with the AVH network [56], was not directly tied to symptom occurrences. The second negative trial referred to peak activation from a motor-based fMRI AVH-capture scan ([54], raising concerns about distinguishing the “response” network from per-hallucinatory activity [27]. A third trial demonstrated resting-state fMRI-based neuronavigation efficacy against sham [57], with 2 and 6-weeks effect-sizes comparable to our MULTIMODHAL study, which compared neuro-guided rTMS to active T3P3 rTMS.

This superiority underscores the transformative potential of our approach. Indeed, the MULTI-MODHAL trial further advances fMRI-guided TMS toward establishing a validated, neurobiologically informed treatment for drug-resistant SCZ. To achieve this goal, it combines two key elements: (i) precision psychiatry (using a phasic brain marker directly linked to AVH occurrences), and (ii) personalized care (neuronavigation to position the rTMS coil at a subject-specific target beyond a one-size-fits-all approach).

Although resting-state functional or structural MRI designs assume that the identified neural traits associated with AVHs are stable over time, our results demonstrate that compared with non-responders, responders exhibit a significantly stronger interaction between the rTMS induced E-field and the phasic AVH network obtained via capture-fMRI, despite no difference in the baseline resting-state FC at the stimulation site. These results support the idea that state-based, neurobiologically defined targets may outperform resting-state proxies or purely anatomical landmarks at the temporo-parietal junction, and that precise targeting matters more for clinical response than the total energy delivered to the brain.

A second strength of the trial lies in its parallel-group superiority design, which better controls for placebo effects, consistently reported in previous sham-controlled rTMS trials of AVHs [58]. Indeed, none of our participants were denied access to active stimulation, thereby allowing for identical: (i) professional attention, (ii) scalp sensations at the stimulation site, and (iii) perceived technological credibility of the neuronavigation device that was similarly deployed in both groups (even if it was only used effectively in the neuro-guided group). The integrity of the blind was confirmed as participants’ guesses about their treatment were no better than chance.

Despite these strengths, several limitations should be acknowledged. First, the relatively modest sample size (35 participants per group) may limit generalizability. However, this is offset by the high levels of daily-life symptoms and impairments observed in the investigated drug-resistant cohort (see Table 1), and the demanding nature of the fMRI symptom-capture procedure, which is only contributive in patients experiencing frequent AVHs. Consequently, recruitment was limited to only two sites, to ensure harmonization of the fMRI-capture procedure. Of course, this may raise the question of real-world scalability of fMRI symptom-capture methods. We recently addressed this by validating an automated fMRI-based AVH decoder [30] that generalizes to out-of-sample patients, enabling target definition and improving clinical practicability, even in centers with limited MR signal-processing expertise.

Future replication studies (notably including those relying on our new automated fMRI-capture procedure [30]) will need to confirm the present results, by recruiting larger cohorts across multiple centers. While MULTIMODHAL focused on the specific impact of neuro-guided rTMS, we believe future work could also consider integrating emerging stimulation paradigms, such as continuous theta-burst (cTBS) to further optimize therapeutic outcomes. Additionally, such replications could benefit from incorporating recent methodological developments to refine coil orientation, utilizing a priori E-field modeling [59], robotization [60] and rTMS adjusted amplitude to the scalp-to-cortex distance of each target [61].

To our knowledge, the MULTIMODHAL study is among the longest follow-up rTMS trials, thereby providing valuable insight into the long-term effects of this technique. However, a potential limitation arises from the progressive withdrawal of participants after 3-months of follow-up, which is partly due to the recurrence of hallucinations to baseline-levels. This scenario increased variance in later measurements, despite the use of multiple imputation methods (including main, planned and unplanned sensitivity analyses). Although the interpretation of long-term effects should be performed with caution in this context, the persistence of a significant difference at 3 months (but not afterward) in the completers analysis (per-protocol Supplementary Table 5) suggests a gradual loss of the therapeutic benefit beyond this time point. Both replication and adaptive studies are now necessary to precisely determine the effect duration, as well as optimal maintenance strategies.

In summary, the MULTIMODHAL trial confirms that neuro-guided 1-Hz rTMS over a brain target defined by using a validated fMRI-capture procedure, is not only effective at relieving drug-resistant AVHs in schizophrenia patients, but also superior to conventional T3P3 stimulation (NNT = 3.5), with comparable tolerance. Future improvements may come from: (i) developing brain-based predictors of response, to help clinicians in better identifying patients who will benefit from such optimized rTMS treatment, and (ii) exploring maintenance stimulation schemes, to extend the effect of rTMS beyond 3-6 months post-treatment.

## MULTIMODHAL Study Group

Hamid Aftisse, Lucie Bailleul, Briac Batailley, Victoire Bénard, Alexandre Bonord, Jacques Catteau, Claire-Lise Charel, Thibault Dablin, Marie-Laure Darcas, Laurent Defromont, Morgane Demeulemeester, Julien Dumont, Amélie Duvaux, Marion Eck, Philippe Fallon, Marie-José Fontaine, Thomas Fovet, Tristan Gaggioli, Axelle Gharib, Isabelle Guesdon, Michel Goudemand, Abdennour Hamek, Mickael Henon, Mathilde Horn, Frédéric Ivanez, Christine Lajugie, Agnès Lambrichts, Bertrand Lavoisy, Perrine Lekadir, Isle Lespoix, Virginie Lopacinski, Francesco Macri, Sophie Mayeur, Caroline Michaut, Sylvain Montalvo, Iulia Nedelescu, Jean-Michel Pailleux, Sylvie Platteau, Alexandre Pouchet, Marianne Ramonet, Claire Rascle, Valentine Riehl, Sylvie Robert, Frédéric Sédivy, Eric Thomazeau, Maxime Tiberghien, Juliette Tiprez, Elisabeth Trolle, Catherine van Nedervelde, Emmanuel Wertz.

## Supplementary Methods

### Description of the motor-free fMRI hallucination capture method

Patients wore headphones and earplugs to attenuate scanner noise and were asked to lie in the scanner in a state of wakeful rest with their eyes closed. They reported hallucinatory episodes immediately after the capture fMRI run using a previously validated post-fMRI interview procedure [19, 20, 62]. This interview allowed us to specify the global frequency of AVHs during the scanning procedure, the relative moment(s) of AVH occurrence and the subjective duration of AVH events. These post-fMRI questionnaires were subsequently used to identify cortex-based ICA components capturing AVHs for each patient (referred to as the “two-step method” in previous validation papers). If no AVHs occurred during the run, a new capture fMRI session was scheduled. Only patients who reported at least one AVH experience while being scanned were included for further analyses.

### Description of the electrical field modeling method

The Electrical field modeling was performed in two steps. First, individualized head models were constructed for each patient using CHARM-TMS [63] provided by SimNIBS, integrating T1-weighted MR images, tissue segmentation and surface reconstruction outputs generated by FreeSurfer. Second, we simulated the electrical field propagation using SimNIBS (Python package) [46] and the abovementioned reconstructed head models, positioning the MagVenture Cool-B65 coil either over the standard T3P3 site or the personalized simulation target.

### Description of the MRI/fMRI preprocessing steps

Resting-state functional connectivity (FC) analyses were performed using the anatomical and functional MRI data in the MNI space of each patient, including T1-weighted scans, BOLD functional images and gray matter (GM) segmentation standard outputs from fMRIPrep [64]. The T1-weighted images were mapped to an atlas-based parcellation (Glasser atlas) to produce the brain regions of interest. The participant-wise rTMS stimulation sites, which were defined as the thresholded E-field maps (at 15% RMT), were used as the primary seed for the FC profile calculated from the resting-state BOLD data for each patient. To identify an optimal number of clusters, a silhouette index was used and k=2 was chosen to apply a spectral K-means clustering algorithm to the resulting connectivity profiles [48, 65]. Afterward, a manual relabeling of these two resulting clusters, for each participant, was performed to ensure consistency across the dataset. Group-level FC maps were separately computed for cluster #1 and cluster #2 for the responders and non-responders. To clarify the similarities/differences in connectivity profiles between responders and non-responders, the top 20 parcels (based on FC strength) common to both clusters were compared.

## Supplementary Figures & Tables

The Methods and Results sections are supplemented by two additional figures (SupFig.4 and 5) and three supplementary tables (SupTab. 4, 5, 6).

**Figure 4.**
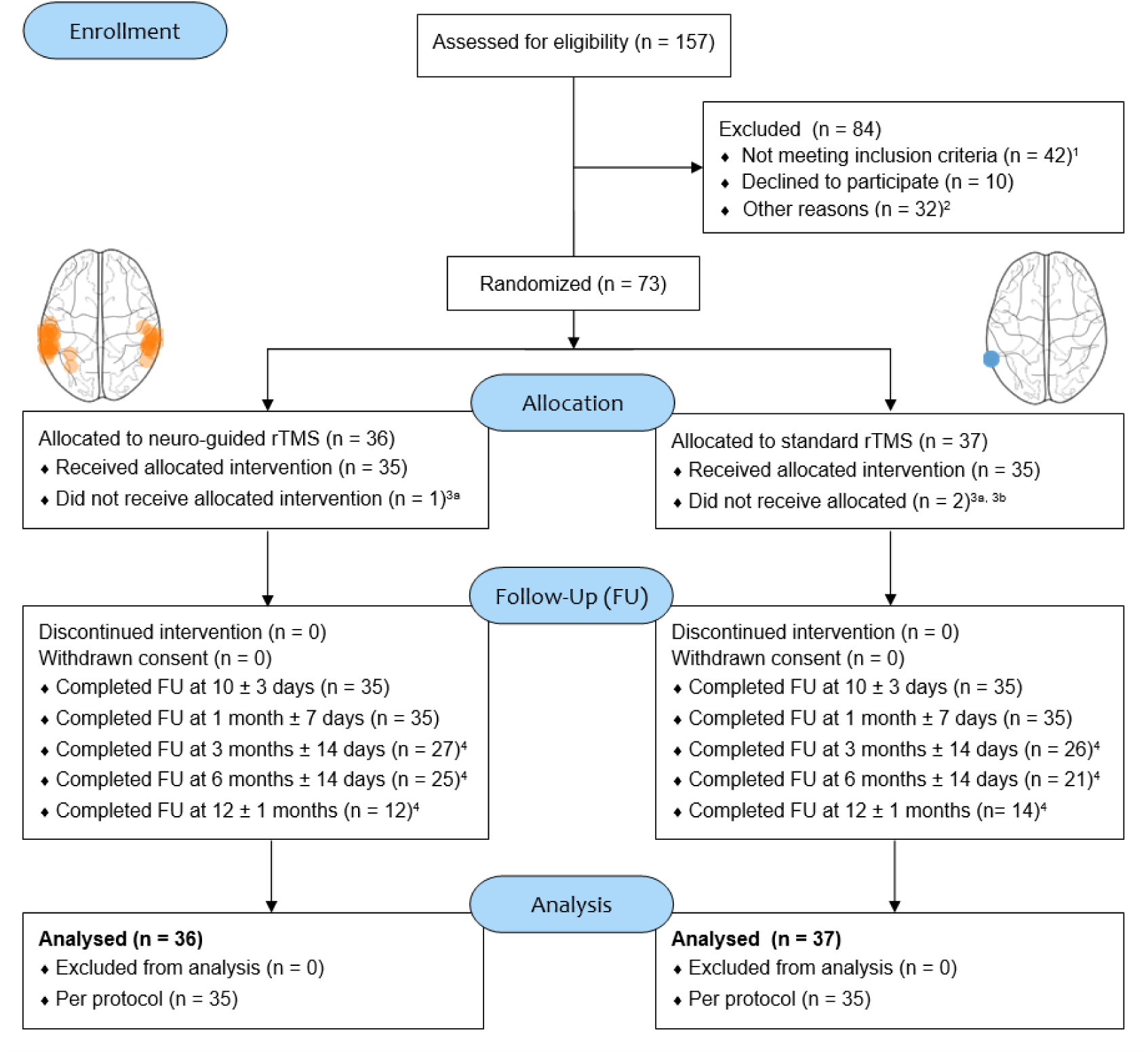
CONSORT flow-diagram of participation in the MULTIMODHAL trial. **(1)** 42 patients did not meet inclusion criteria because of: a revised diagnosis after screening (n=14), claustrophobia (n=1), insufficient or remitted hallucinations at the screening visit (n=12), rTMS contraindication (n=1), severe addiction (n=1), liberty deprivation context (n=1), ongoing pharmacological change with treatment not yet stabilized (n=4), inability to provide informed consent (n=3). **(2)** 23 patients were excluded from the trial due to: transport phobia, relocation to another country and alternative therapeutic choice. Three patients were lost to follow-up, due to: **(3a)** two consecutive non-contributing MRI (n=2); **(3b)** trial leave after one rTMS session (n=1); Finally **(4)** 44 patients progressively left the trial after hallucinations severity returned to their basal level.

**Figure 5.**
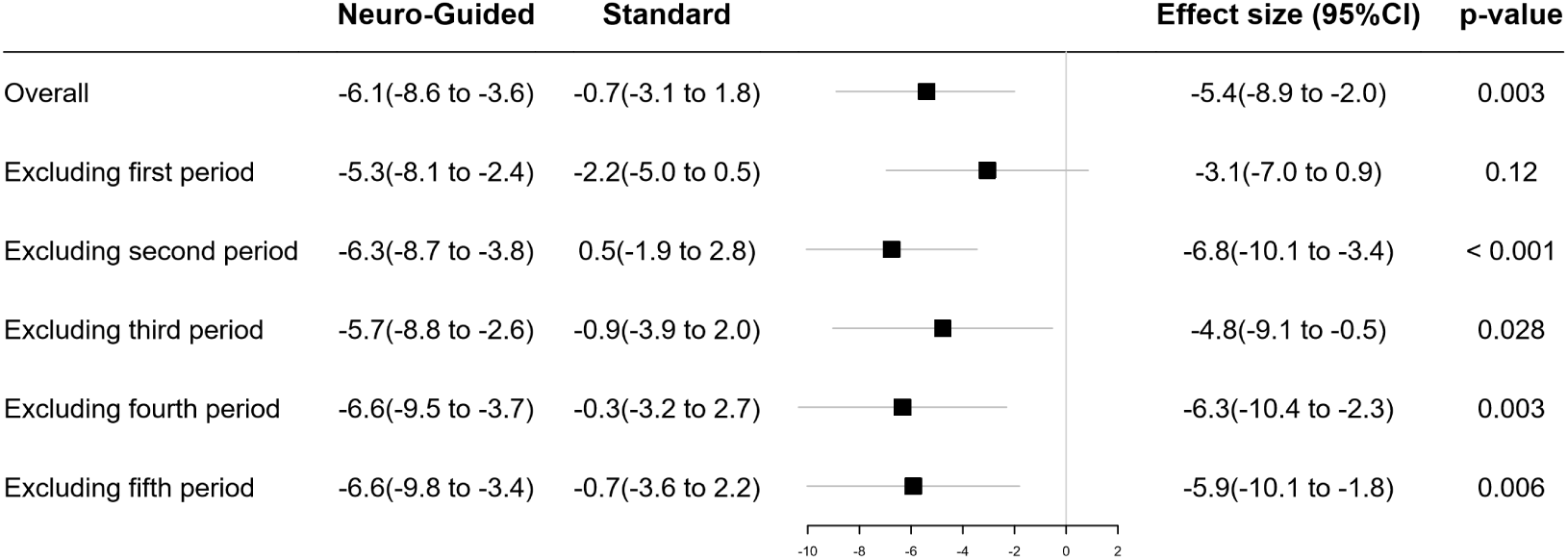
Leave-one-out sensitivity analysis of the primary outcome (Auditory Hallucination Rating Scale [AHRS] score change at 1 month) over the 2011 - 2022 inclusion period. The forest plot displays the overall treatment effect alongside sequential sub-period exclusions (excluding the first through fifth periods chronologically). For each analysis, values are presented as: (i) mean changes (with 95% confidence intervals, 95%CI) for the neuro-guided and standard groups, (ii) the resulting between-group effect size, and (iii) the corresponding p-value. These findings demonstrate that the rTMS treatment effect remains consistent and robust across all iterations, with the exception of the model omitting the first sub-period (non-significant, despite a similar trend), thereby limiting potential confounding from shifts that may have occurred in clinical practice over time.

**Table 4.** Primary and Secondary Outcomes in Sensitivity Analyses after handling Missing Data by Multiple Imputations.

|  | Neuro-Guided<br>(n=36) | Standard<br>(n=37) | Effect Size (95% CI) | p-Value |
| --- | --- | --- | --- | --- |
| **Primary Outcome (AHRS Score)** |  |  |  |  |
| 1-0 diff. | -6.0 (-8.6 to -3.5) | -0.5 (-3.0 to 2.1) | -5.5 (-8.9 to -2.1) | **0.002** |
| 3-0 diff. | -4.3 (-7.2 to -1.3) | -0.6 (-3.4 to 2.2) | -3.6 (-7.5 to 0.4) | **0.075** |
| 6-0 diff. | -6.9 (-10.1 to -3.6) | -3.7 (-7.1 to -0.2) | -3.2 (-8.0 to 1.7) | 0.19 |
| 12-0 diff. | -8.0 (-16.4 to 0.5) | -5.7 (-10.1 to -1.3) | -2.3 (-10.2 to 5.8) | 0.57 |
| **Secondary Outcomes** |  |  |  |  |
| PANSS total score 1-0 diff. | -13.5 (-20.1 to -6.9) | -8.9 (-15.6 to -2.3) | -4.5 (-13.7 to 4.8) | 0.34 |
| PANSS positive sub-score 1-0 diff. | -2.5 (-3.5 to -1.5) | -0.8 (-1.7 to 0.3) | -1.7 (-3.0 to -0.4) | **0.013** |
| PANSS P3-item 1-0 diff. | 0 (-2 to 0)<sup>a</sup> | 0 (-1 to 0)<sup>a</sup> | -0.4 (-0.7 to -0.1)<sup>b</sup> | **0.019** |
| VAS frequency 1-0 diff. | -1 (-3 to 0)<sup>a</sup> | 0 (-1 to 0)<sup>a</sup> | -0.5 (-1.0 to -0.1)<sup>b</sup> | **0.016** |
| VAS intensity 1-0 diff. | -1.9 (-2.6 to -1.2) | -0.3 (-1.0 to 0.4) | -1.6 (-1.7 to -1.5) | **0.001** |
| GAF score 1-0 diff. | 3.7 (0.3 to 7.1) | 0.6 (-2.6 to 3.8) | 3.1 (-1.3 to 7.7) | 0.17 |
| CGI score at 1 Month | 3 (3 to 4)<sup>a</sup> | 4 (3 to 4)<sup>a</sup> | -0.3 (-0.8 to 0.2)<sup>b</sup> | 0.19 |

<sup>a</sup> Median (25th to 75th percentile). <sup>b</sup> Standard mean difference on rank-transformed data.
Values are presented as least-squares means (95% CI) derived from cLDA models, unless otherwise specified. Effect sizes represent mean differences (95% CI) in baseline-adjusted changes from baseline, except where noted. AHRS stands for *Auditory Hallucination Rating Scale*. PANSS stands for *Positive And Negative Syndrome Scale*. VAS stands for *Visual Analogue Scale*. GAF stands for *Global Assessment of Functioning* scale. CGI stands for *Clinical Global Impression* scale.

**Table 5.** Primary and Secondary Outcomes in Sensitivity Analyses restricted to the Per-Protocol population.

|  | N | Neuro-Guided<br>(n=28) | N | Standard<br>(n=30) | Effect Size (95% CI) | p-Value |
| --- | --- | --- | --- | --- | --- | --- |
| **Primary Outcome (AHRs Score)** |  |  |  |  |  |  |
| 0 (baseline) | 28 | 26 ±7 | 30 | 29 ±6 |  |  |
| 1 Month | 28 | 19 ±9 | 30 | 27 ±8 |  |  |
| 3 Months | 20 | 20 ±9 | 19 | 26 ±9 |  |  |
| 6 Months | 18 | 19 ±10 | 15 | 24 ±8 |  |  |
| 12 Months | 9 | 17 ±11 | 10 | 23 ±6 |  |  |
| 1-0 diff. |  | -6.9 (-9.6 to -4.2) |  | -1.2 (-3.85 to 1.51) | -5.72 (-9.54 to -1.90) | **0.004** |
| 3-0 diff. |  | -5.8 (-8.9 to -2.7) |  | -0.1 (-3.2 to 3.0) | -5.7 (-10.0 to -1.3) | **0.013** |
| 6-0 diff. |  | -6.6 (-10.0 to -3.1) |  | -1.8 (-5.4 to 1.8) | -4.7 (-9.7 to 0.4) | 0.065 |
| 12-0 diff. |  | -8.0 (-14.1 to -1.9) |  | -2.7 (-8.5 to 3.2) | -5.3 (-13.7 to 3.2) | 0.20 |
| **Secondary Outcomes** |  |  |  |  |  |  |
| PANSS total score |  |  |  |  |  |  |
| 0 (baseline) | 28 | 80 ±24 | 30 | 76 ±14 |  |  |
| 1 Month | 28 | 66 ±25 | 29 | 69 ±20 |  |  |
| 1-0 diff. |  | -13.6 (-21.0 to -6.2) |  | -6.9 (-14.1 to 0.5) | -6.8 (-17.0 to 3.5) | 0.19 |
| PANSS positive sub-score |  |  |  |  |  |  |
| 0 (baseline) | 28 | 17 ±5 | 30 | 16 ±4 |  |  |
| 1 Month | 27 | 14.2 (12.8 to 15.6) | 29 | 16.2 (14.8 to 17.7) |  |  |
| 1-0 diff. |  | -2.6 (-3.7 to -1.5) |  | -0.58 (-1.6 to 0.6) | -2.0 (-3.5 to -0.5) | **0.011** |
| PANSS P3-item |  |  |  |  |  |  |
| 0 (baseline) | 28 | 5 (5 to 6)<sup>a</sup> | 30 | 6 (5 to 6)<sup>a</sup> |  |  |
| 1 Month | 27 | 5 (4 to 5)<sup>a</sup> | 29 | 5 (5 to 6)<sup>a</sup> |  |  |
| 1-0 diff. |  | -1 (-2 to 0)<sup>a</sup> |  | 0 (-1 to 0)<sup>a</sup> | -0.5 (-1.0 to -0.1)<sup>b</sup> | **0.036** |

Table 5 – continued from previous page
|  | N | Neuro-Guided<br>(n=28) | N | Standard<br>(n=30) | Effect Size (95% CI) | p-Value |
| --- | --- | --- | --- | --- | --- | --- |
| VAS frequency |  |  |  |  |  |  |
| 0 (baseline) | 28 | 5 (3 to 8)<sup>a</sup> | 30 | 8 (5 to 9)<sup>a</sup> |  |  |
| 1 Month | 28 | 4.3 (2 to 5)<sup>a</sup> | 30 | 6.6 (4 to 9)<sup>a</sup> |  |  |
| 1–0 diff. |  | –1 (–5 to 0)<sup>a</sup> |  | 0 (–2 to 0)<sup>a</sup> | –0.7 (–1.1 to –0.2)<sup>b</sup> | **0.009** |
| VAS intensity |  |  |  |  |  |  |
| 0 (baseline) | 28 | 5.8 ±2.7 | 30 | 7.2 ±2.0 |  |  |
| 1 Month | 28 | 4.3 ±2.7 | 30 | 6.6 ±2.4 |  |  |
| 1–0 diff. |  | –1.7 (–2.5 to –1.0) |  | –0.3 (–1.0 to 0.6) | –1.5 (–2.5 to –0.4) | **0.008** |
| GAF score |  |  |  |  |  |  |
| 0 (baseline) | 28 | 44 ±14 | 30 | 46 ±14 |  |  |
| 1 Month | 28 | 48 ±13 | 30 | 46 ±14 |  |  |
| 1–0 diff. |  | 4.2 (0.2 to 8.1) |  | 0.7 (–3.0 to 4.5) | 3.5 (–1.7 to 8.8) | 0.19 |
| CGI score at 1 Month | 28 | 3 (3 to 4)<sup>a</sup> | 30 | 4 (3 to 4)<sup>a</sup> | –0.3 (–0.8 to 0.2)<sup>b</sup> | 0.22 |

**Notes:** <sup>a</sup> Median (25th to 75th percentile). <sup>b</sup> Standard mean difference on rank-transformed data.
Values are presented as least-squares means (95% CI) derived from cLDA models, unless otherwise specified. Effect sizes represent mean differences (95% CI) in baseline-adjusted changes from baseline, except where noted. AHRs stands for *Auditory Hallucination Rating Scale*. PANSS stands for *Positive And Negative Syndrome Scale*. VAS stands for *Visual Analogue Scale*. GAF stands for *Global Assessment of Functioning* scale. CGI stands for *Clinical Global Impression* scale.

**Table 6.** Unplanned Sensitivity Analysis for Longitudinal Analysis of Primary Outcome Measure.

|  | Neuro-Guided<br>(n=36) | Standard<br>(n=37) | Effect Size (95% CI) | p-Value |
| --- | --- | --- | --- | --- |
| Primary Outcome (AHRS Score) |  |  |  |  |
| 1-0 diff. | -5.5 (-7.7 to -3.2) | -0.6 (-2.8 to 1.7) | -4.9 (-8.0 to -1.7) | 0.003 |
| 3-0 diff. | -4.6 (-6.7 to -2.5) | -0.2 (-2.3 to 2.0) | -4.4 (-7.4 to -1.5) | 0.005 |
| 6-0 diff. | -5.5 (-7.7 to -3.3) | -1.4 (-3.6 to 0.9) | -4.1 (-7.2 to -0.9) | 0.013 |
| 12-0 diff. | -4.8 (-7.0 to -2.7) | -1.0 (-3.1 to 1.2) | -3.8 (-6.9 to -0.8) | 0.015 |

Values are presented as least-squares means (95% CI) derived from cLDA models, unless otherwise specified. Effect sizes represent mean differences (95% CI) in baseline-adjusted changes from baseline, except where noted. AHRS stands for *Auditory Hallucination Rating Scale*.

## Data availability

The data supporting the findings of this study will be available from the corresponding author after publication.

## Author Contributions

The project was supervised by RJ (conceptualization, project administration and funding acquisition). Data were collected and prepared by DR. Data were analyzed by JL, EC, ZA, PD, PY and RJ. All the authors contributed to the data interpretation and writing of the paper.

## Acknowledgement

This preprint was created using the LaPreprint template.

